# Accelerometer Data Harmonization and Feature Engineering for Gait Analytics

**DOI:** 10.64898/2026.11.05.26367336

**Authors:** F. Ph. Carrle, F. Schmidt, A. Reichenbach

## Abstract

Gait analytics is utilized for a wide range of health-related applications, ranging from fitness tracking to medical diagnostics, and beyond. Inertial measurement units that are ubiquitously available in mobile devices provide an accessible means of gait analysis but differences in sensor specifications, placement, and recording protocols limit comparability across studies. This study developed and evaluated an analysis-centered data warehouse and standardized quality-assessed processing pipeline to facilitate the integration and joint analysis of heterogeneous gait accelerometer data. Six publicly available datasets including a total of 935 persons were integrated into a star-schema data warehouse, and feature comparability was assessed across datasets and sensor locations. Gender classification served as a proof- of-concept downstream analysis. Gait-cycle extraction, which serves as foundation for feature extraction, was successfully achieved with an accuracy above 99% for the dataset that provides ground truth information. Comparability varied across datasets and sensor configurations, with correlation coefficients between average gait cycles ranging from 0.5 to close to 1.0. Centrally positioned sensors generally show greater correspondence than peripheral sensors, although differences in statistical features persist even between sensors at nominally identical locations. Gender classification performance remains largely near chance level, and no consistent benefit from pooling datasets could be established. These findings demonstrate that heterogeneous gait accelerometer data can be integrated and processed within a common analytical framework, but standardized processing alone does not ensure comparability of the resulting features. The proposed open-source framework implemented in Python provides a foundation for reproducible cross-study gait analysis and for investigating residual heterogeneity associated with datasets and sensor configurations.

**Author Summary:** Sensors in smartphones and wearables such as smartwatches and fitness trackers enable background recording of human walking behaviour in everyday life. This data can be used in a range of health-related applications and research, both in the fitness and in the medical domain. In order to develop robust analytical methods and applications, large and diverse datasets are desirable. However, data recorded for such purposes often differ in sensor placement, hardware specification, recording procedures, and data structure, impeding the compilation of composed datasets. In this study, we develop a harmonization framework that organizes and processes accelerometer-based gait data in a standardized way. This includes a database structure, automated gait-cycle extraction, quality-control procedures, and feature-extraction methods that can be readily applied across multiple source datasets. Using six publicly available datasets, we demonstrate the technical feasibility of our approach. A subsequent downstream analysis highlights the possibilities and limits of this approach and demonstrates where differences between datasets cannot be harmonized but need to be taken into account in subsequent analyses. By providing the framework as an open-source resource, we aim to facilitate future efforts to unlock the power of large gait datasets and therewith support reproducible gait research.

## Introduction

Quantification of the human gait is of interest to a variety of disciplines. In the wider context of health, gait features can on the medical side support the diagnostic procedure and monitoring of disorder or therapeutic progress [1] in orthopaedics [1–3], neurology (e.g., Alzheimer’s and Parkinson’s disease [4, 5] or stroke [6]), and psychiatry (e.g., schizophrenia, anxiety, depression, and other mood disorder [7–10]). On the performance side, fitness tracking for well-being purposes [11–13] as well as analytics in recreational and competitive sports are increasingly utilizing gait tracking and similar measurements [14]. Unrelated to health applications, gait features are harnessed in further use cases such as biometric authentication [15–18]. The gold standard for gait analytics has long been data from motion capture systems, force plates, and similar laboratory equipment [19, 20]. Those systems are challenged by mobile sensors such as inertial measurement units (IMU) comprising accelerometers and gyroscopes [19, 21, 22]. Those sensors are versatile, lightweight, easily available, cheap, and ubiquitous due to their integration in standard smartphones [23] and other smart devices. Their usefulness in gait analytics has been demonstrated by several studies showing in general a good correspondence between characteristics extracted from video- or force sensor-based methods and from smartphone IMU data [14, 24, 25]. The characteristics assessed in those studies are predominantly spatio-temporal, i.e., step cycle features such as step length, step time, or cadence [24], or additionally kinematic features [14].

Depending on the use case, different features are extracted from the raw gait time series. They include spatial, temporal, spatio-temporal, kinematic, and kinetic features [14, 19]. Besides general changes in spatio-temporal features such as slowing or reduction of step length [5], specific changes in kinematic features such as smoothness, or the symmetry in spatio-temporal feature are characteristics of interest in a wide range of health applications [6]. Gait asymmetry in particular has been found to be associated with balance instability at older age [26–28], compromised mental health status [29], and is also prevalent in individuals diagnosed with motor-related neurological disorders such as Parkinson’s disease [4, 30–33], post-stroke conditions [34–37], and Multiple Sclerosis [38–40]. Given its clinical relevance, the reduction of gait asymmetry is a targeted outcome in therapeutic interventions for those disorders [41].

Challenges for application of IMU data in gait analysis and related decision support systems cumulate in the lack of robustness and generalisability of the features and models used for those purposes [5, 14, 20]. Diversity in sensor specifications, recording setups, sensor locations [24, 42], data quality, and study designs [20] hamper the comparability and replication of research findings [43]. On top of those variations stemming from recording conditions is the lack of standardized format for data storage and processing [20, 44], which leads to laborious data harmonization efforts whenever data from different studies or simply from different sensors needs to be processed in a comparable fashion. Gait analytics can be improved by leveraging large and diverse datasets [20, 44], and with replication of results with demarcable, and if possible, publicly available, datasets [45]. The demand for such datasets and study designs is aggravated by the application of increasingly complex and therefore data-greedy algorithms for analysis purposes, especially in the light of increasingly widespread use of deep learning [5, 46–48], and the demand for translating research findings into practise [20, 44]. The possibility to collect large research datasets with IMUs is easy to realize yet storage, processing, and comparison of the data is still laborious due to the diversity on several ends.

This study presents 1) an analysis-centered data scheme (data warehouse (DWH) with star schema [49–51]) for accelerometer data from gait tasks including participant- and study-specific additional data, 2) a quality-assessed data processing pipeline for data harmonization, pre-processing, feature extraction, and analytics, and 3) a proof-of-concept analysis on the combined dataset. The quality of the gait cycle extraction algorithm is assessed with a quality metric and additionally evaluated against ground truth for one of the datasets included. As feature sets, we provide 1) statistical features from the gait cycles including asymmetry features and 2) the normalized full gait cycle from the Euclidean norm of the jerk. For evaluation purposes, the features were systematically compared across body sides, sensor locations, and datasets. Furthermore, the feature sets were subjected to exemplary analyses on gender classification with different configurations of included datasets. Data scheme and extract-transform-load (ETL) processes are based on the data of six publicly available datasets comprising gait accelerometer data from both standalone IMUs and sensors built into smartphones [52–58]. However, data scheme and processing pipeline are designed to be extendable to incorporate further data formats, additional information, and further processing steps such as additional data quality checks or additional features. Both technical artifacts are provided in an open-source toolbox with accompanying technical documentation.

## Results

### Data Storage

The data is stored in a star schema comprised of central fact tables each containing specific parts of recorded data or their derivates. Accelerometer data is stored raw, preprocessed, in gait cycles, and as statistical features. Additional fact tables contain raw electromyography (EMG), gyroscope, and magnetometer data. The dimension tables connected to the fact tables contain additional information that provide the analytical context for the fact data, such as participant characteristics, or sensor positions and specifications.

### Quality Management

We verified the gait cycle extraction process with the Marea [52] dataset that provides a ground truth of the gait cycles based on force sensitive resistors. The average dominant peak detection accuracy for the left and right ankle sensor was 99.3±2.1% and 99.9±0.2%, respectively. The heel strike is recorded 56.7±18.6ms and 58.9±65.4ms before the detected peak across all data samples for the left and right foot, respectively.

In order to evaluate the data quality independent of a ground truth, the quality of the extracted gait cycles (n=343,431) from the accelerometer data was assessed by correlating each individual uniform cycle with the corresponding average cycle. In order to be included in further analyses, we set a threshold of 0.7. 255,542 cycles, i.e., 74.4% of the cycles, fulfilled this criterion.

### Statistical Evaluation of Harmonized Data

In order to pool data across datasets or sensors, the features used for analyses need to be comparable. We first evaluated comparisons of exemplary sensors where the data stem either from different datasets, sensor locations, or body sides for illustrative purposes. The comparison between two sensors on the right waist (same location) from the datasets OU-ISIR-SAID [55] and Vajdi [53] (different datasets) shows only a moderate overlap in the average gait cycle (*r*=0.82; Fig 1, top). Along this line, most of their statistical features were not comparable (Fig 1, bottom).

**Fig 1.**
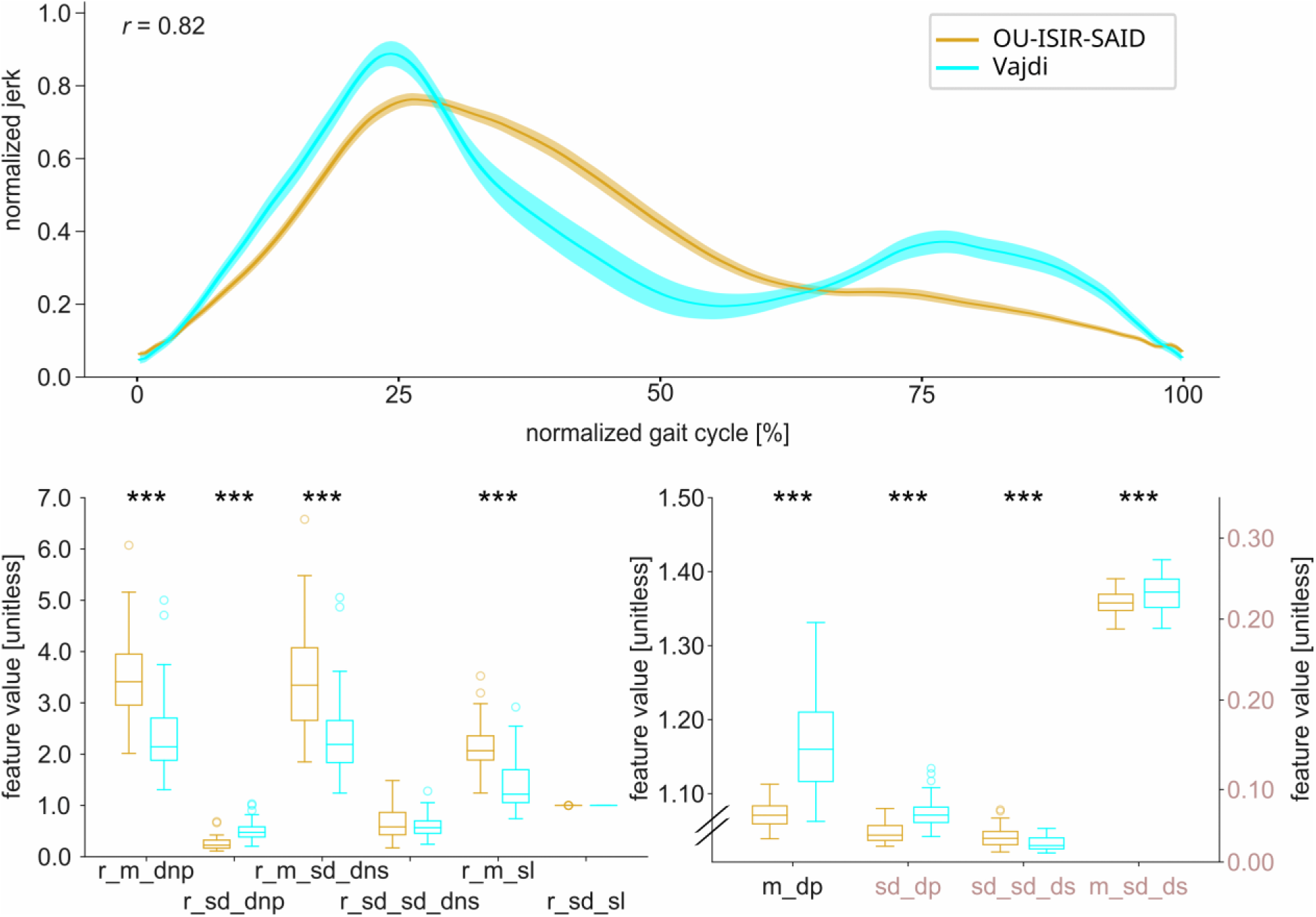
Comparison of uniform gait cycles (top) and statistical features (bottom) of the sensors on the right waist between the OU-ISIR-SAID [55] and Vajdi [53] datasets. The gait cycles are temporally normalized and the statistical features are divided between ratio features for gait asymmetry quantification (left) and measures of central tendency and dispersion (right). *** p<0.001, uncorrected. Shaded bars denote 95% confidence interval and outliers in the boxplots are defined as 1.5 x the interquartile range below or above the first or third quartile, respectively. Abbreviations: r_m_dnp = ratio of mean values of dominant and non-dominant peaks, r_sd_dnp = ratio of standard deviations of dominant and non-dominant peaks, r_m_sd_dns: ratio of mean standard deviations of dominant and non-dominant steps, r_sd_sd_dns: ratio of standard deviations of standard deviations of dominant and nondominant steps, r_m_sl: ratio of means of dominant and non-dominant step length, r_sd_sl: ratio of standard deviations of dominant and non-dominant step length, m_dp: mean value of dominant peaks, sd_dp: standard deviation of dominant peaks, sd_sd_ds: standard deviation of standard deviations of dominant steps, m_sd_ds: mean standard deviation of dominant steps

In contrast, the pairing of two other datasets (Vajdi [53] and DBHuGa [56]) even with similar but different sensor locations (left front pocket *vs*. left thigh) shows better harmonization results. The average gait cycles correlate higher (*r*=0.96; Fig 2 top) and the differences between the statistical features were slightly less although still prevalent (Fig 2, bottom).

**Fig 2.**
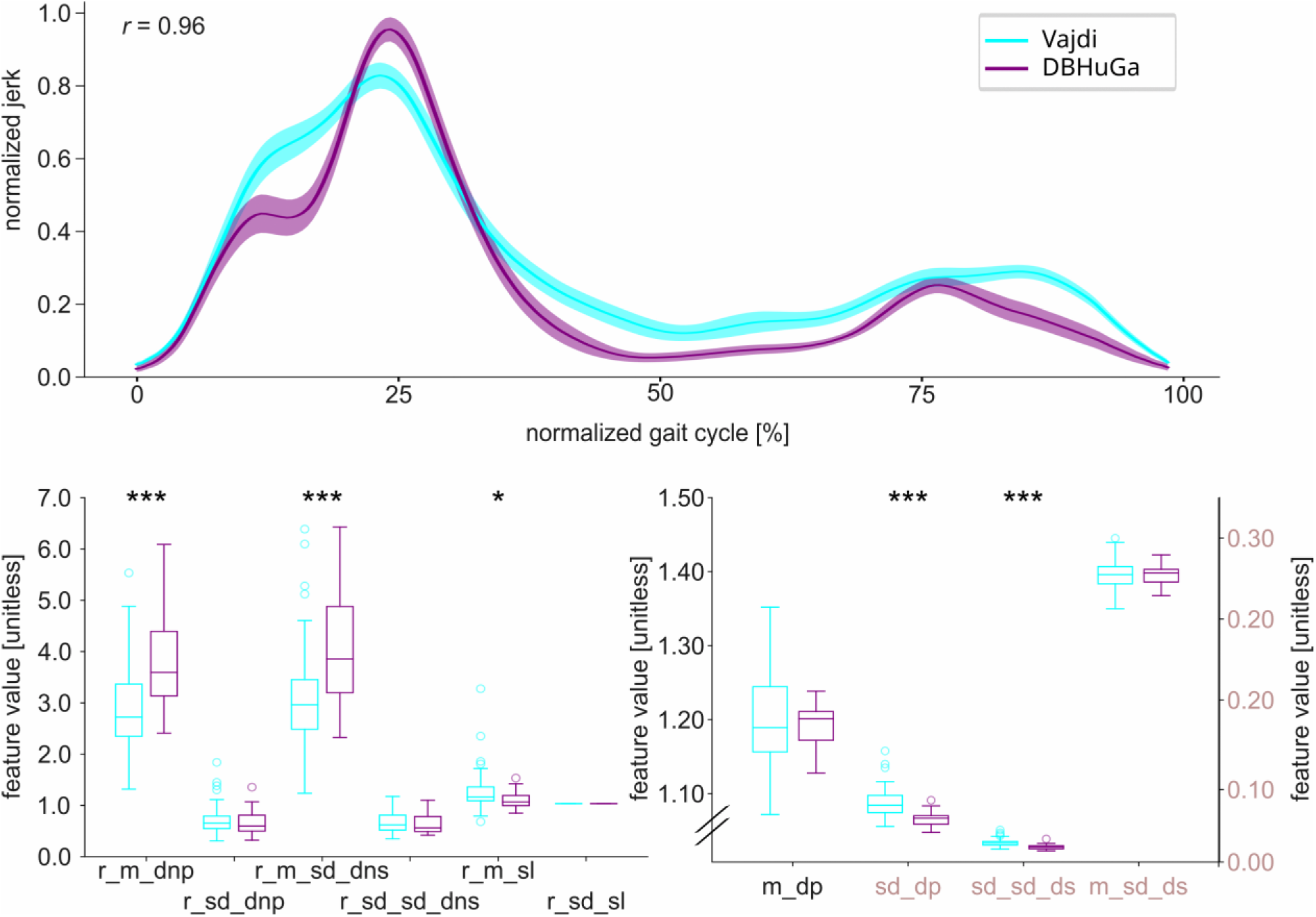
Comparison of uniform gait cycles (top) and statistical features (bottom) of the left front pocket sensor from the Vajdi [53] and the left thigh sensor from the DBHuGa [56] dataset. Presentation conventions and abbreviations analogue to Fig 1; * p<0.05, *** p<0,001, uncorrected.

There is a very good correspondence for sensors of the same dataset in the same location but for opposite body sides (DBHuGa [56] shin (Fig S1) and OU-ISIR-SAID [55] waist (S6)), decreasing for sensors with increasingly different locations (DBHuGa [56] comparison of left shin to left thigh (Fig. S3) and left wrist (S5)).

A less detailed but more comprehensive picture emerges from the full correlation matrix between all sensors in all datasets (Fig 3). The sensor data from the center waist electrodes correlate very highly independent of their dataset of origin (Fig 3, small triangle). Waist sensor placed on one side of the body follow closely regarding their correlation strength. We find a general tendency for the sensor placed on the far end of the extremities (ankle and wrist) to have a lower correlation with those placed more towards the bodies center of weight. The sensors from the OU-ISIR datasets have a visibly lower correlation with sensors from other datasets then the rest of the comparisons (Fig 3).

**Fig 3.**
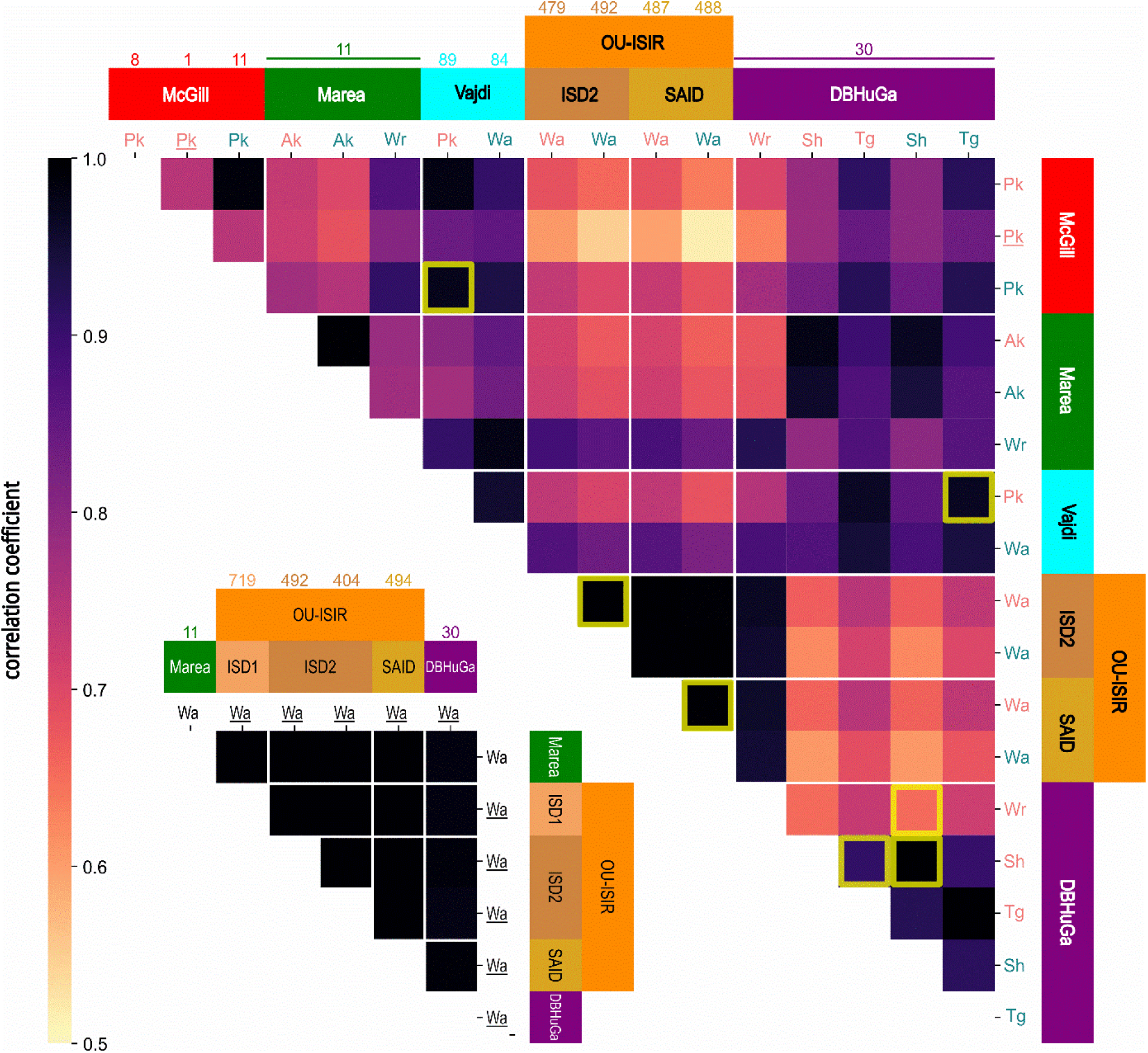
Correlations between the average uniform gait cycles of each dataset and sensor position combination. Results are split into centered sensors (small matrix) and non-centered ones (big matrix) because the two phases of a gait cycle are more evenly distributed for the centered sensors. The exemplary comparison combinations used for the detailed analyses have an olive border. Reddish colours indicate the sensor is placed on the left side of the body, blueish colours for right and black for center. The underline indicates the sensor being placed on the back. The number indicates the number of participants that have recorded data for that particular sensor position. Abbreviations: Pk = Pocket, Ak = Ankle, Wr = Wrist, Wa = Waist, Sh = Shin, Tg = Thigh

In order to put the correspondence across sensor positions and datasets in perspective and assess the variability of gait within participants, we also calculated each sensor’s within and across subject correlations (Fig 4). The across subject correlations of the McGill [57], Marea [59], and DBHuGa [56] sensors are in a similar order as the higher correlations across sensors and databases and consistently higher than the within subject correlations. The across subject correlations in the Vajdi [53] and OU-ISIR datasets [54, 55] are markedly lower and in the range of the within subject variability.

**Fig 4.**
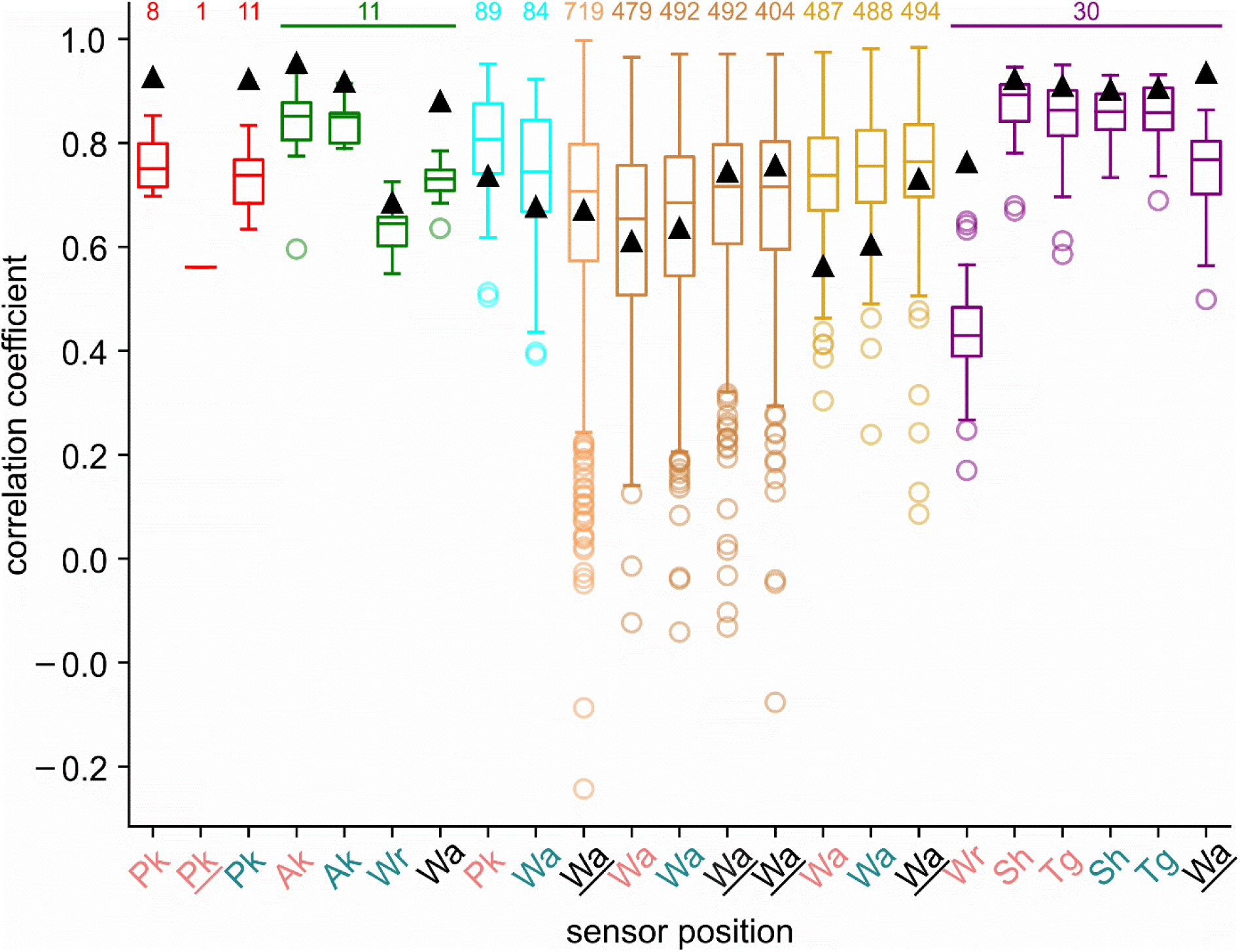
Across and within subject correlations of uniform gait cycles. The black triangles denote the average correlation coefficient between the average cycles of each individual participant (across subject correlations). The boxplots represent the distribution of the average correlation coefficients for each individual participants uniform cycles (within subject correlations). The numbers on top of the boxes indicate the number of subjects per boxplot Colour and font coding of the datasets and sensors analogue to Fig 3. Only cycles that meet the threshold of 0.7 were used here.

### Gender Classification on Harmonized Data

The ultimate test for harmonized data is its utilization in a downstream classification task, comparing performance of individual datasets with joint performance as well as generalization across datasets. We chose a gender classification task since this information is available in all but one datasets and the gender of a person can be detected in its gait [60, 61]. In order to match the dimensionality across feature sets we condensed the 200 time points of the unified gait cycle with a principal components analysis (PCA) to the ten first components that explain a variance of 75.2±11.1%.

Using again the two sensors on the right waist (same location) from the datasets OU-ISIR-SAID [55] and Vajdi [53] (different datasets) (cf. Fig 1) we have to conclude that the features we used for this task do not seem well suited for gender discrimination, at least not from the right waist sensors of these two datasets (Fig 5). Most of the classifiers cannot discriminate the gender and neither a feature set nor a classification algorithm or a training setup has a distinct advantage over the other (all *p*>.05).

**Fig 5.**
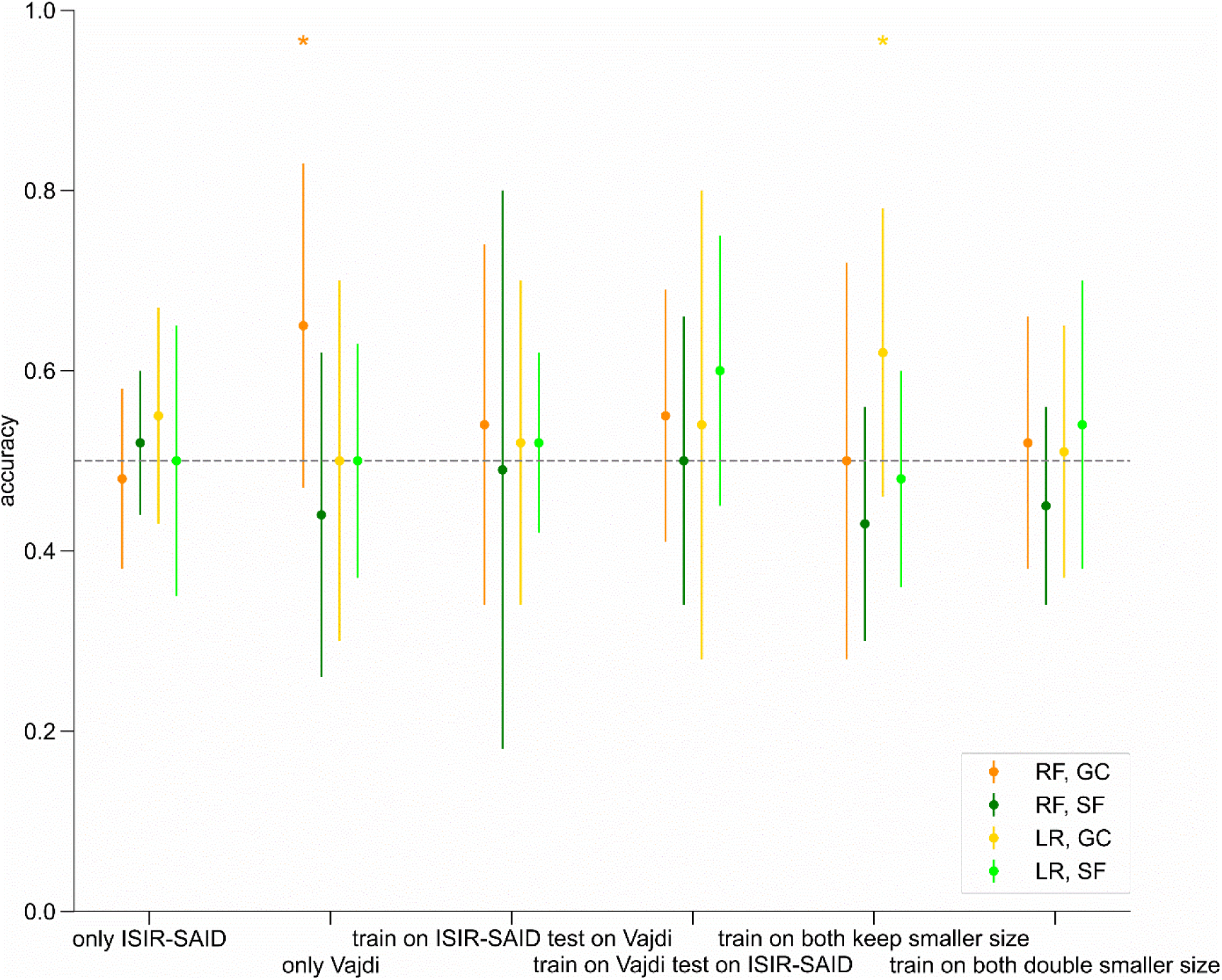
Accuracies for gender-classification with the sensors on the right waist for the OU-ISIR-SAID [55] and Vajdi [53] datasets. The six training setups were each tested with two feature sets and two classification algorithms. Error bars denote the STD of the classification accuracy across the 8 folds. * p<0.05, uncorrected. Abbreviations: RF = random forest, LR = logistic regression, GC = gait cycles, SF = statistical features

The features from sensors in the left front pocket and left thigh, respectively, from the Vajdi [53] and DBHuGa [56] datasets (cf. Fig 2) seem to be similarly inadequate for gender classification (**Fig 6**). However, here the statistical features tend to perform better in general (*F_1,96_*=15.052, *p*<0.001) albeit with marked differences across training setups (*F_5,96_*=2.416,*p* =0.041).

**Fig 6.**
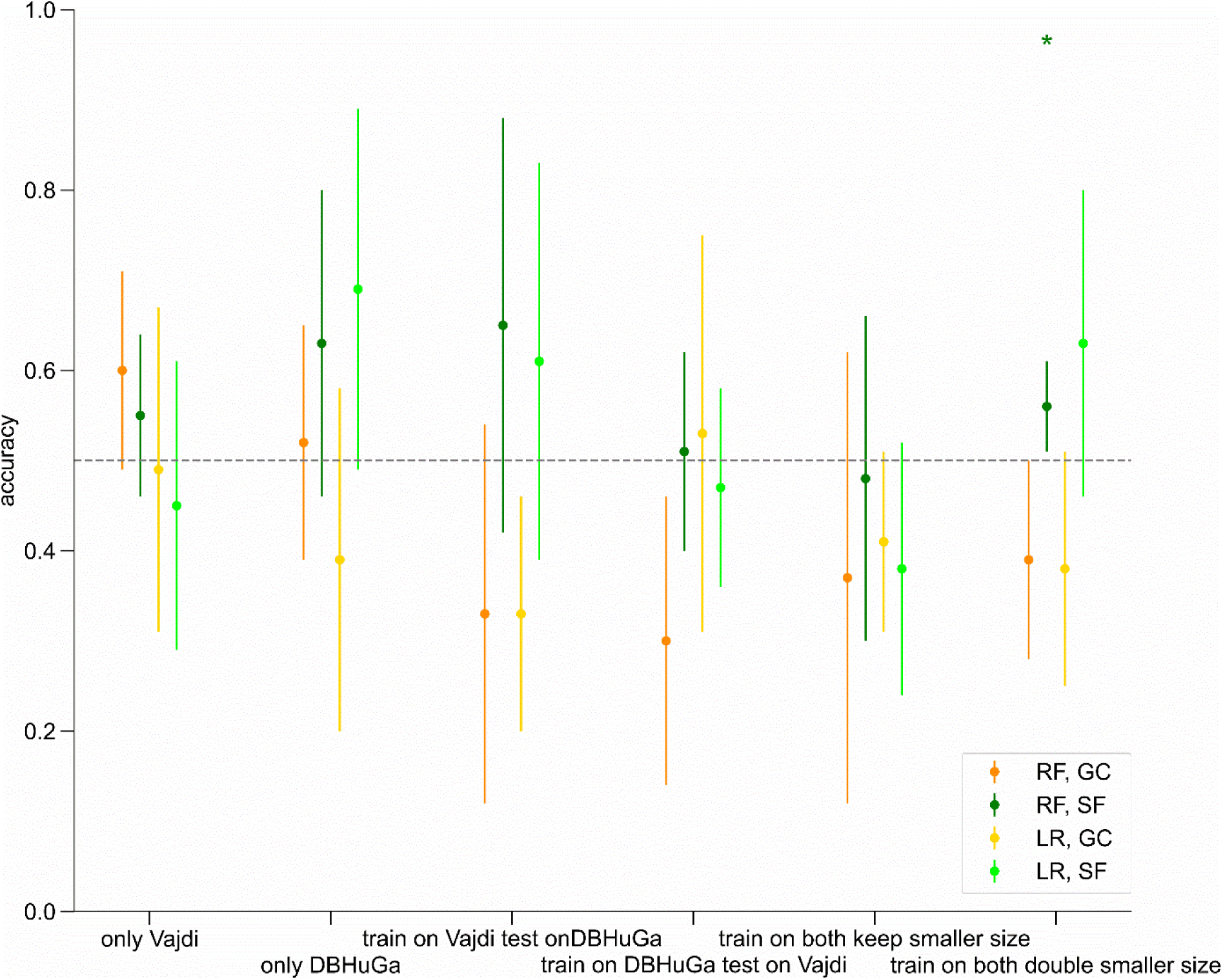
Accuracies for gender-classification with the sensors in the left front pocket from the Vajdi [53] and on the left thigh from the DBHuGa [56] dataset. The six training setups were each tested with two feature sets and two classification algorithms. Error bars denote the STD of the classification accuracy across the 5 folds. * p<0.05, uncorrected. Abbreviations: RF = random forest, LR = logistic regression, GC = gait cycles, SF = statistical features

The classification results based on the remaining sensor pairings (Figs. S2, S4 and S7) present a similar picture. Given the overall around chance level classification performance, an advantage of harmonizing and pooling the data can neither be confirmed nor refuted.

## Discussion

This study developed and evaluated an analysis-centered data warehouse and processing pipeline for harmonizing heterogeneous accelerometer data acquired during walking. The technical framework integrates participant-, study-, sensor-, and recording-specific information with standardized processing steps for gait-cycle extraction, quality assessment, and feature generation. Across six publicly available datasets, the results demonstrate that such a framework can substantially reduce technical heterogeneity and enable joint analysis of data originating from different studies. At the same time, the analyses show that technical harmonization does not necessarily result in comparable resulting gait representations. Residual differences related to dataset, sensor location, and body side remain evident, both for normalized gait cycles and for derived statistical features. This distinction between harmonization of data processing and comparability of the underlying measurements presents important implications for cross-study gait analysis.

The results from the statistical evaluation demonstrate that data can be represented in a common analytical format but still lack analytical comparability. For example, accelerometer recordings obtained from the right waist in the OU-ISIR-SAID [55] and Vajdi [53] datasets were processed using the same pipeline and nominally represent the same sensor location, yet the resulting average gait cycles show only moderate correspondence, while several statistical features differ substantially. This finding indicates that differences between studies cannot be attributed solely to data format or preprocessing. Rather, sensor characteristics, recording protocols, participant populations, and other study-specific factors may remain embedded in the resulting features even after standardized processing. Therefore, technical harmonization is a necessary first step for creating a common representation in which residual heterogeneity can be quantified and investigated, rather than a procedure that completely removes heterogeneity. This issue is well known in other modalities, e.g., batch effects in genomics data [62, 63] or scanner effects in magnetic resonance imaging (MRI) [64–68]. Adjustment methods such as Combating Batch (ComBat, [62]), originally developed for genetic batch effects but transferred successfully to features derived from a variety of MRI modalities[64–67], provide tools for aligning the distribution characteristics of the features while preserving differences in participant characteristics of interest. While those approaches may be suitable for the statistical features derived from the accelerometer data, the partially distinct forms of the unified gait cycles suggest that this feature might not be fit for combination across datasets. These considerations are particularly relevant for multi-study analyses, where apparently standardized inputs may still contain systematic study- or device-specific information.

The results furthermore suggest that harmonization strategies have to explicitly retain sensor location information as an important characteristic of the signal. Data recorded at different body locations are influenced by different biomechanical components of body motion [69, 70] and are therefore not expected to exhibit identical temporal or statistical characteristics even after normalization. Sensors mounted at the waist or other central locations capture a more global representation of the whole-body movement because they are located closer to the body’s center of mass, whereas peripheral sensors are more strongly influenced by limb-specific motion and attachment characteristics [71–73]. Our correlation analysis indicates generally stronger correspondence among centrally positioned sensors, even across datasets, whereas sensors located toward the extremities, such as the ankle and wrist, tend to show lower correspondence with sensors positioned closer to the body center or with sensors from other datasets. This renders the former especially suitable for analyses across datasets. However, depending on the use case, the former or latter is better suited to capture the characteristic of interest [72].

The pairwise comparisons demonstrate that some sensor-position combinations have substantially stronger correspondence, indicating that harmonization can facilitate meaningful comparisons under appropriate conditions. The across- and within-subject analyses provide additional evidence that the magnitude of variability depends on both the dataset and the sensor configuration. In several datasets, across-subject correlations were higher than the corresponding within-subject variability, whereas other datasets showed the opposite pattern. This heterogeneity suggests that the relative contributions of subject-specific gait characteristics and recording-specific factors are not constant across datasets. Consequently, pooled analyses should account for the individual structure of the data and only opt for a combination of datasets when the conditions are met.

The gait-cycle extraction procedure provides an important foundation for our harmonization approach. When evaluated against annotated gait events in the MAREA dataset [52], dominant-peak detection showed high accuracy for both left and right ankle sensors, with mean accuracies over 99%. These findings support the feasibility of extracting a consistent gait-cycle representation directly from single sensor accelerometer recordings, even when explicit gait-cycle annotations are unavailable. However, this validation should be interpreted within the scope of the available ground truth. The algorithm was evaluated against event annotations from a single dataset and only for ankle-mounted sensors; therefore, these results do not establish equivalent performance across all sensor positions represented in the data warehouse. This limitation is particularly relevant for peripheral and non-peripheral sensors, for which the relationship between local acceleration patterns and conventional gait events may differ. Future validation using independently annotated datasets and multiple sensor locations would therefore be important to establish the generalizability of the extraction algorithm. Along this line, we decided for a unified processing of the data from each sensor individually instead of a global gait cycle extraction across sensors because of the heterogeneity in number of sensors and their locations across datasets. An extension of the preprocessing pipeline might be useful for specific sets of sensors when a global gait cycle with temporally aligned sensor cycles is necessary for addressing specific research questions.

The quality-control procedure offers a complementary safeguard against unreliable cycle extraction. Of the 343,431 extracted gait cycles, 74.4% met the predefined correlation threshold of 0.7 with the corresponding mean gait cycle and were retained for downstream analysis. This demonstrates the practical value of an automated quality-assessment step when integrating data from heterogeneous sources. At the same time, the finding that approximately one quarter of all extracted cycles did not meet the criterion indicates that the choice of quality threshold has a potentially substantial effect on the resulting analytical dataset. The threshold of 0.7 was selected for the present analysis and is configurable in the pipeline. Nevertheless, the current study does not establish whether this threshold is optimal across datasets, sensor locations, or gait conditions. The within-subject correlations demonstrate substantial differences for sensor positions and datasets suggesting that it might be recommended to adapt individual thresholds. Importantly, however, a low correlation with the mean cycle is not necessarily equivalent to poor data quality, as atypical but genuine gait cycles may also differ from the recording-specific mean. Especially in clinical populations, this variability can even be an important feature of interest [74–76].

The rational for the gender-classification analysis was to provide a proof of concept for the use of the harmonized data in downstream analysis, rather than to develop an optimized gender-classification model. Classification performance was generally close to chance for the examined sensors, and a consistent benefit from combining data across datasets or sensor locations could therefore not be established. These findings do not demonstrate that the harmonization framework is ineffective. Rather, they illustrate that successful technical harmonization does not automatically translate into improved performance in a downstream machine-learning task. Furthermore, the configuration of the six training setups provides a suitable strategy for proper evaluation of joint data usage. Comparing the power of the increased data size achieved by pooling with the performance of the individual datasets and the pooled data while keeping dataset size provides a comprehensive test for improvement via data pooling. Training on one dataset and testing on the other provides a test unbiased to preprocessing for the generalizability across studies.

The strength of this study is the use of multiple publicly available datasets with substantial variation in participant numbers, recording duration, sensor type, sensor location, and study characteristics. This heterogeneity reflects the conditions under which a general harmonization framework ultimately needs to operate. The data warehouse additionally retains contextual information alongside the sensor measurements, providing a basis for investigating these sources of variation rather than discarding them during preprocessing. The framework is also designed to accommodate additional processing steps, sensor modalities and further datasets, while the technical artifacts are provided in an open-source toolbox with accompanying documentation. These endeavours support reproducibility and provide a basis for extending the framework beyond the datasets evaluated here.

Some limitations, nevertheless, need to be considered. First, the available datasets differed substantially in participant characteristics, recording duration, sensor configuration, and study protocol, and these factors are often intrinsically confounded. For example, an observed dataset effect cannot necessarily be attributed to the dataset itself because it may reflect differences in hardware, placement, participant population, or recording conditions. Second, several potentially informative comparisons could not be performed because the required metadata or features were unavailable. Consequently, the pairwise comparisons should be regarded as representative examples rather than a complete characterization of all possible sources of heterogeneity. Future studies might explicitly model contextual factors such as age, footwear, walking surface, and other recording conditions in order to disentangle their contributions [77].

Overall, the present study demonstrates that heterogeneous accelerometer datasets can be integrated into a common analytical framework with standardized gait-cycle extraction, quality management, and feature engineering. At the same time, the results show that technical harmonization alone does not remove systematic differences associated with datasets and sensor configurations. The principal contribution of the framework is therefore to provide a reproducible infrastructure in which the differences across datasets can be identified, quantified, and incorporated into subsequent analyses. Such an approach facilitates more transparent cross-study comparisons, reproducible feature engineering, and ultimately more robust development and validation of gait-analysis methods across heterogeneous real-world datasets.

## Material and Methods

This chapter provides an overview about the data for the initial load of the DWH, the concept for data storage and data processing as well as details about feature engineering and data evaluation. Technical details of the data scheme, the processing steps, and data quality management is provided as separate technical documentation with the supplementary material. An open-source code repository (https://github.com/ReichenbachLabHHN/GaitDWH) provides all technical artifacts to generate the database scheme and the processing pipeline.

### Data and Participants

Six publicly available datasets (**Table 1**) from a variety of countries are included in the initial data scheme and processing pipeline. All datasets include accelerometer data from walking, which is used in this study. Possible additional sensor data is only included in the initial data loading process, but the remainder of the data scheme and processing pipeline can be easily extended to include information from this data as well.

**Table 1.**
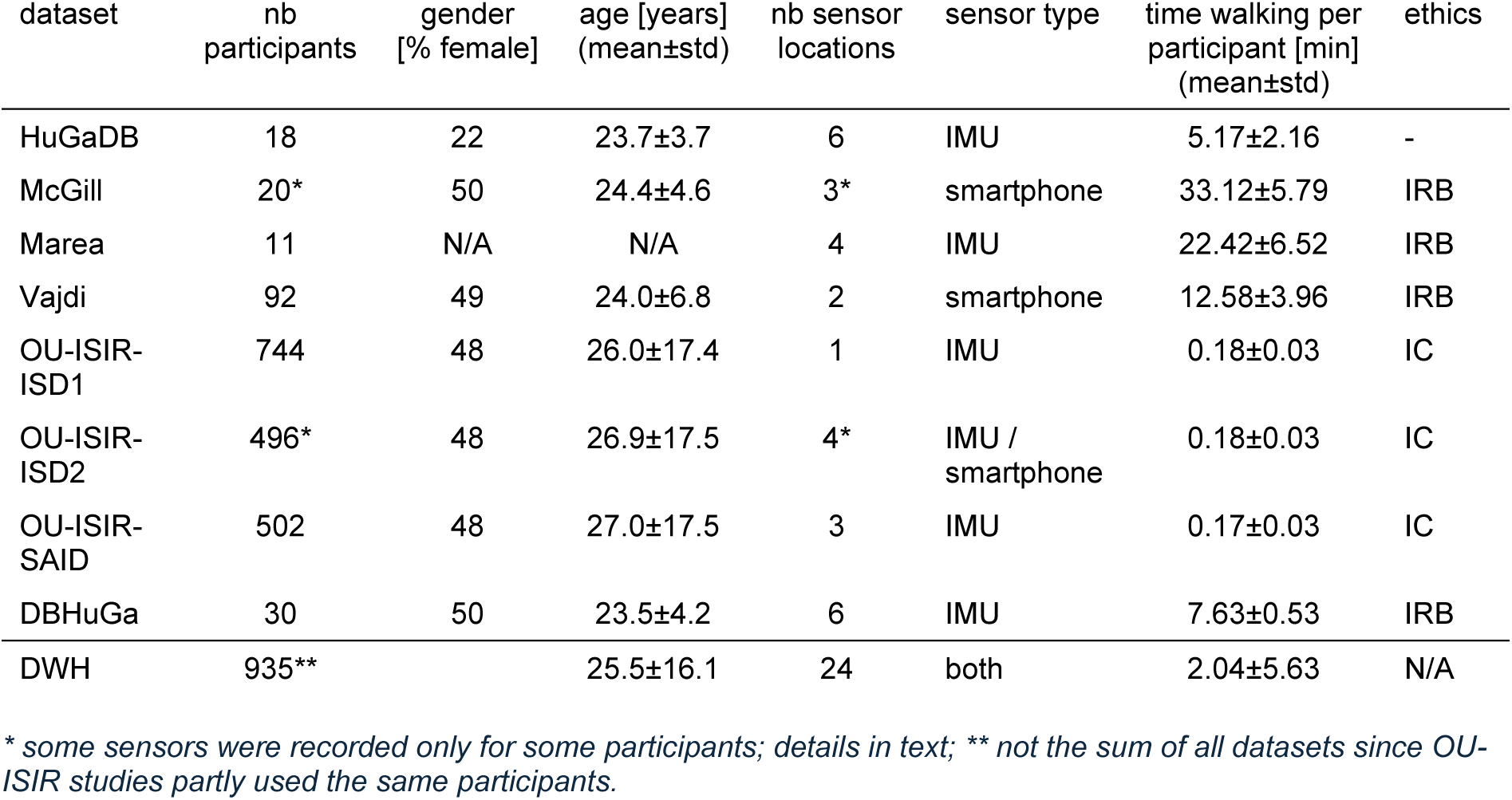
Characteristics of the original datasets and the harmonized data warehouse. Sensor locations for each dataset are depicted in *Fig. 7*. Abbreviations: datasets see text; IMU: inertial measurement unit; IRB: institutional review board; IC: informed consent

The Human Gait Database **(HuGaDB)** [58] from Russia additionally includes recordings of eleven non-walking activities and provides gyroscope measurements and electromyography data as well as accelerometer data. Since this dataset does not provide information about ethics protocols for data collection, this data is not used for analysis purposes in this study. The McGill University Gait Dataset **(McGill)** [78] from Canada uses only one of the three available sensor locations for each participant, and furthermore includes magnetometer data. The Movement Analysis in Real-world Environments using Accelerometers Gait Database **(MAREA)** [52] from Sweden includes not only walking but also running activity performed under varying conditions. It provides ground truth annotations generated via Force Sensitive Resistors (FSRs) placed on both feet, capturing the frame-level occurrence of four gait-cycle events. The Vajdi Gait Database **(Vajdi)** [53] from the USA additionally contains gyroscope data. The OU-ISIR Gait Database from Japan represents a large collection of gait related datasets. Two subsets are particularly relevant for this work: the Inertial Sensor Dataset **(OU-ISIR-ISD)** [54] and the Similar Action Inertial Dataset **(OU-ISIR-SAID)** [55]. Although derived largely from the same set of recordings, these datasets differ with respect to the sensors employed and the specific data used. The OU-ISIR-ISD dataset is partitioned into two subsets. The first (**OU-ISIR-ISD1**) maximizes participant count but uses only one of the four available sensors. The second (**OU-ISIR-ISD2**) includes all four sensors but does not contain recordings for all participants. Both subsets provide gyroscope data in addition to the accelerometer data. Importantly, the two subsets partially overlap in terms of participants and recordings, which must be taken into account when jointly analysing them. The OU-ISIR-SAID dataset contains five different activities, includes gyroscope measurements, and provides a ground-truth annotation in the form of step counts. The Database of Human Gait Performance (**DBHuGa**) [56] from the USA also contains activities beyond walking, and each of its sensors records gyroscope as well as magnetometer data. In total, the selected data sources contain recordings of up to 935 participants from 24 sensors, some of which share similar sensor location (**Fig 7**) [79].

**Fig 7.**
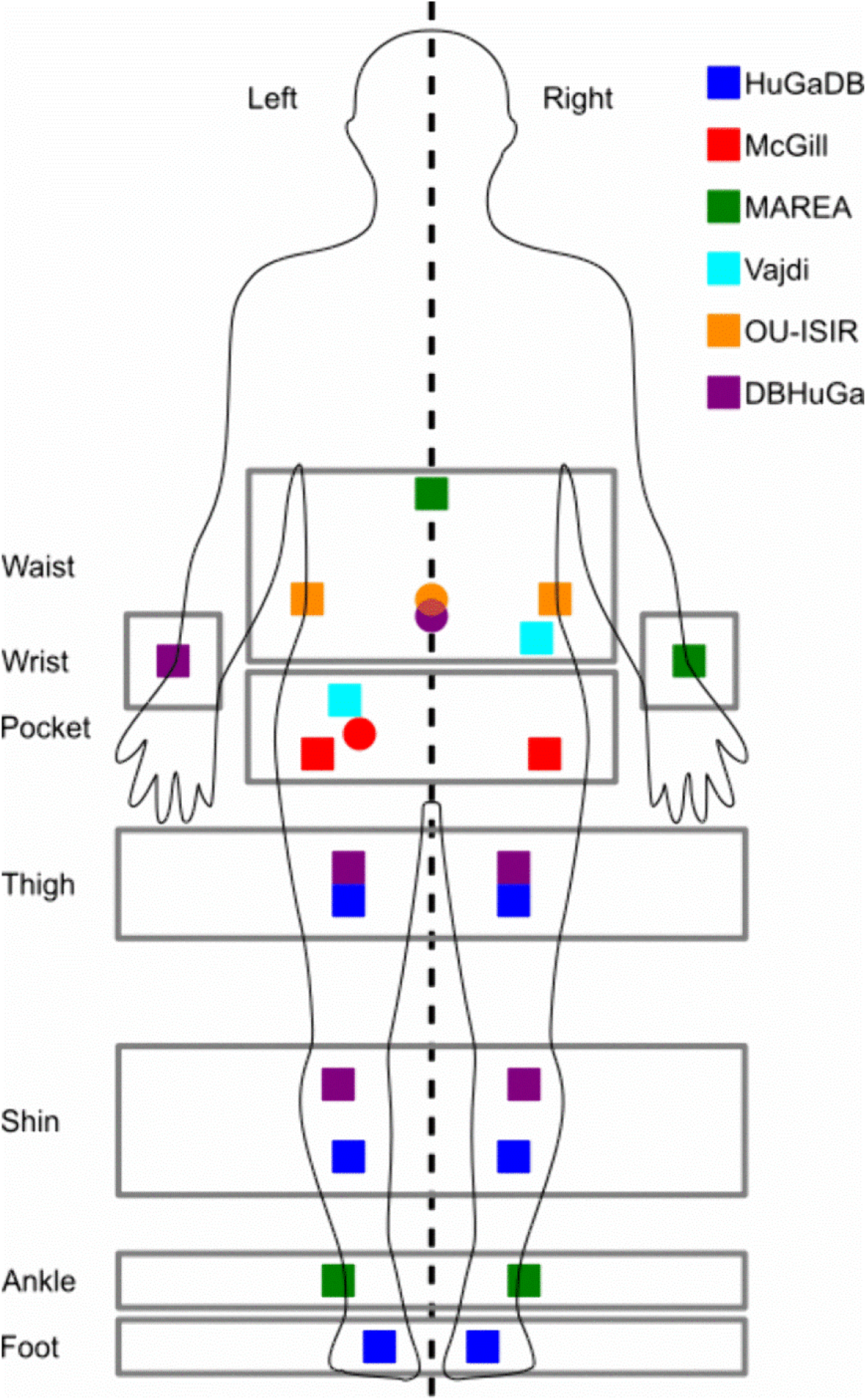
Overview of sensor positions for all datasets. Rectangle indicates a sensor being placed on the front, a circle on the back of the participant. Depicted is a front view of the participant. Abbreviations of the datasets see text; Human outline has been designed using resources from Flaticon.com[79].

The full dataset includes 485 male (age 24.53±16.04 years) and 439 female (age 26.56±16.05 years) participants. Additionally, eleven participants from the MAREA database which does not provide information about the gender.

### Data Scheme

A star-schema–based data model was selected to support the analysis of the data (Fig 8), as this data scheme effectively supports and facilitates large-scale analytical queries [49]. In this structure, one or more central fact tables store the primary observational data while multiple dimension tables provide contextual information relevant to those observations. Each dimension table is directly associated with one or more fact tables, and the dimensions are independent from each other. In contrast to normalized relational designs, the star schema intentionally permits a certain degree of data redundancy to simplify query operations. By reducing the number of required joins, this approach improves computational efficiency at data read-out and is therefore well suited for analytical environments that demand rapid processing of large and complex datasets.

**Fig 8.**
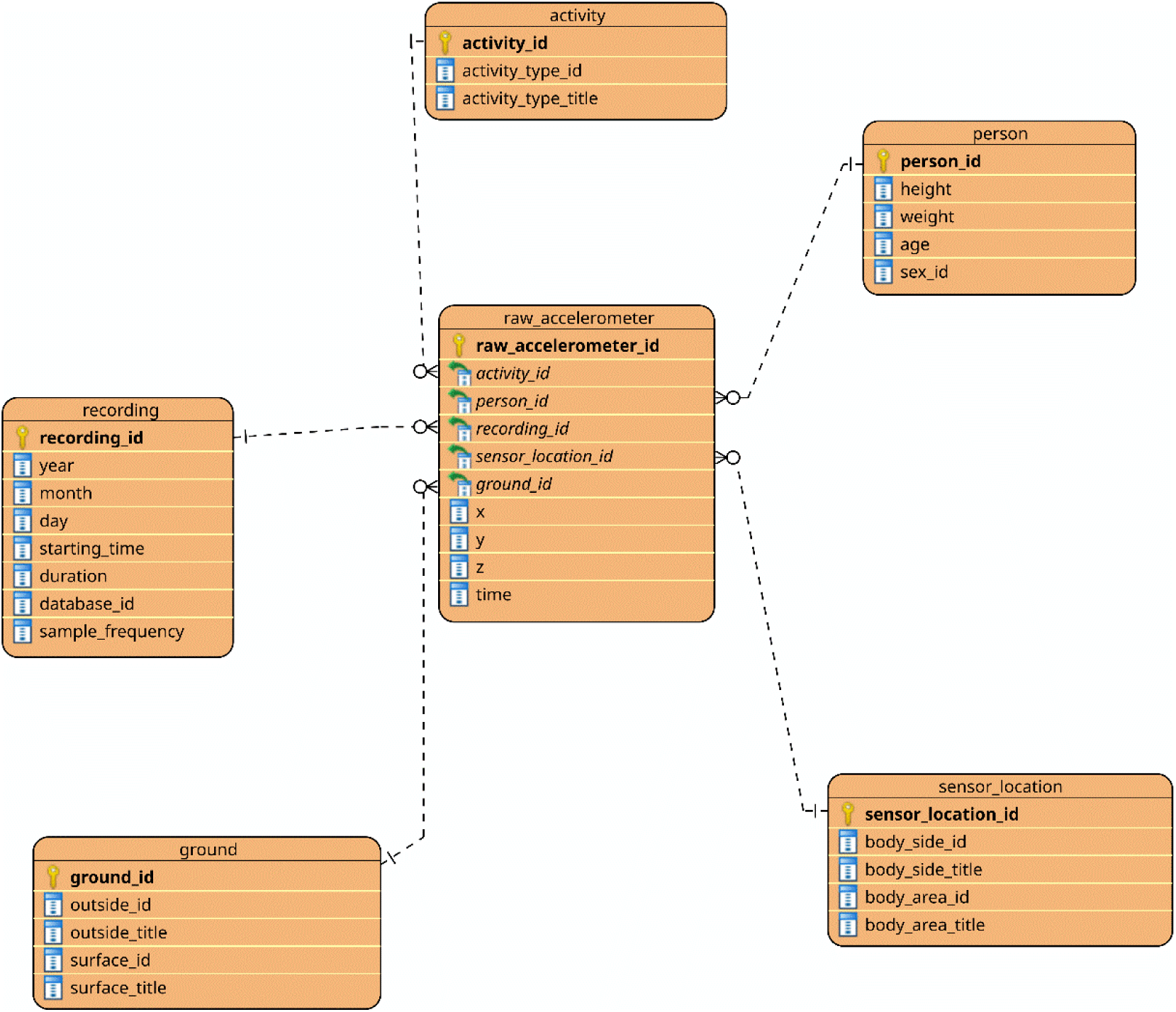
Exemplary star schema for the raw accelerometer data. Note that the surrounding dimension tables are shortened for clarity; only a subset of their columns is included, also some dimension tables are missing for the same reason. The figure is intended to support conceptual understanding of the star scheme structure. Additional fact tables, such as preprocessed accelerometer or extracted features, can share the same dimensions. Figure created with the Visual Paradigm Modeling Tool (Visual Paradigm, Hong Kong, China).

The implemented scheme includes four initial fact tables: Raw Accelerometer, Raw Gyroscope, Raw Magnetometer, and Raw EMG. Only the raw accelerometer data is further processed. They store the respective raw sensor readings together with timestamps to maintain temporal order. Additional fact tables, introduced during feature extraction, are the preprocessed accelerometer data, extracted gait cycles in different normalization steps as well as statistical features extracted from the gait cycles. Eight dimension tables provide complementary information on activities (activity), ground conditions (ground), participants (person), recording metadata (recording), technical sensor information (sensor), sensor placement (sensor_location), step events (step_event), and rotation information for harmonization of sensors (world_coordinate_system).

A comprehensive description of the data model, including a detailed data dictionary, is available in the accompanying technical documentation. SQL scripts for setting up the data scheme with a postgreSQL (The PostgreSQL Global Development Group) database are provided in the code repository.

### Processing Pipeline

#### Individual Extract-Transform-Load Process

The initial ETL process incorporates the full set of available sensor data, resulting in a raw dataset that includes accelerometer measurements from walking and non-walking activities as well as data from additional sensor modalities. Depending on the specific research objectives, the processing pipeline may be extended to accommodate further activity types, facilitated by the accompanying data dictionary.

The data itself is not included as part of the code package and needs to be obtained independently since some of the datasets are only available upon request from the researchers who recorded the data. Moreover, the data warehouse is designed to easily integrate additional recordings, enabling researchers to enrich the repository with their own data if required.

#### Cycle Extraction

The analysis focusses on gait cycles derived from the accelerometer data during walking. Because most available datasets do not explicitly provide gait cycle annotations, we designed and tested an algorithm to extract gait cycles directly from the sensor signals.

Each accelerometer provides tri-axial measurements (*_x_*, *_y_*, and *_z_* components), which inherently depend on the sensor’s orientation, as each axis captures acceleration along a specific spatial direction. To mitigate this orientation dependency and obtain a signal representation that reflects only the underlying dynamic behaviour, the Euclidean norm of the jerk is used for further processing.

To estimate the global step frequency, we identified the first prominent peak in the frequency spectrum. For sensor placements near the body’s center of mass, this peak typically appears around 2 Hz, whereas peripheral sensor locations yield frequencies closer to 1 Hz.

For non-centered sensor placements, a gait cycle is defined as the concatenation of the swing phase of the limb bearing the sensor, resulting in a dominant peak, followed by the contralateral swing phase, which yields a non-dominant peak. Dominant peaks are first identified by locating local maxima that exceed the global mean, while enforcing a minimum inter-peak distance determined by the estimated step frequency. To suppress non-dominant events among these candidates, we apply an amplitude threshold derived from the mean of the candidate peak amplitudes, again subject to the same distance constraint. Assuming an alternating dominant/non-dominant structure, we subsequently examine the inter-peak intervals with respect to the step period to infer and insert missing dominant peaks whose amplitudes may fall below the mean-based threshold.

Non-dominant peaks are then identified by segmenting the signal between successive dominant peaks under the assumption that each segment contains exactly one non-dominant event. Because this event lies between two flanking minima, it is designated as the maximum bounded by the pair of ‘major minima’ (Fig 9). To determine these minima, we first detect all local minima within the segment, excluding those within a small neighbourhood of dominant peaks to avoid false detections. The absolute minimum in the segment is taken as the first major minimum. For each remaining candidate minimum, we compute the highest signal value along the path connecting it to the absolute minimum. Minima that lie in the same valley as the absolute minimum yield lower connecting maxima, whereas those positioned across the opposing valley share the amplitude of the non-dominant peak. Among the minima belonging to this opposing valley, the one with the lowest value is selected as the second major minimum. The non-dominant peak is then identified as the highest local maximum located between these two major minima. The gait cycle is then defined as the major minimum preceding the dominant peak until the major minimum preceding the next dominant peak.

**Fig 9.**
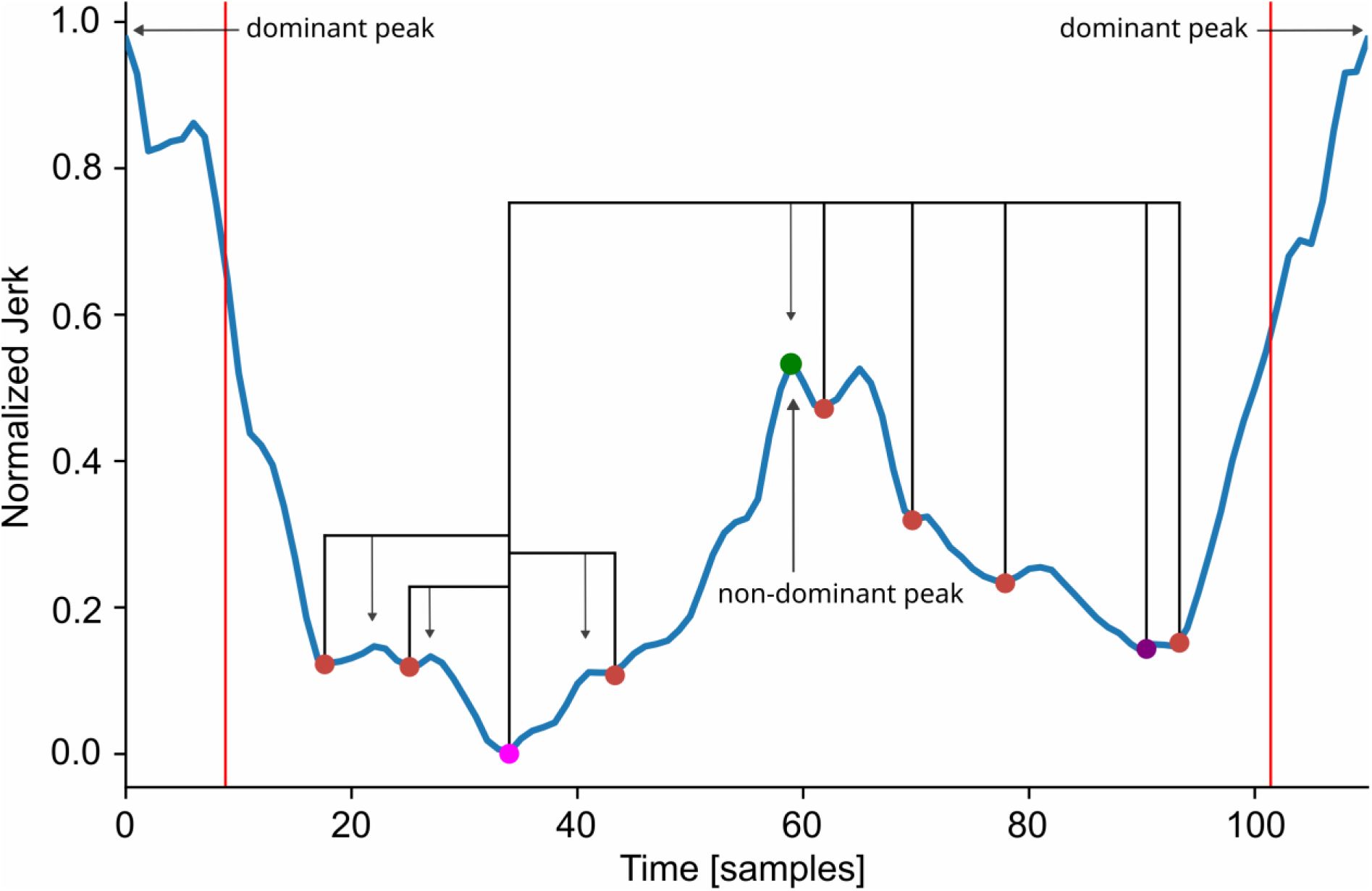
Major minima detection for non-dominant peak detection. All detected minima are marked with a circle. The first minimum is simply the lowest value (hot pink). The second minimum is not the second lowest value since this can be on the same side of the non-dominant peak. The algorithm described in the text finds the true second minimum (dark purple).

For centrally mounted sensors, the extraction of individual gait cycles is accomplished by segmenting the signal at every second minimum between successive peaks. This approach is feasible because contributions from the left and right leg appear with comparable

prominence in the signal, resulting in a symmetric pattern that allows reliable cycle delineation.

#### Ground Truth

To evaluate the performance of the cycle extraction algorithm, we used the ankle-mounted sensors from the MAREA dataset, which provide annotated step-event labels. Heel-strike events are expected to correspond, with a temporal offset, to the dominant and non-dominant peaks identified within the extracted gait cycles. However, because the dataset occasionally contains misordered event annotations, only walking sequences with correctly ordered step-event labels are included in the analysis. Wrist-mounted accelerometer data are excluded, as the dominant peak observed in wrist motion during a gait cycle is not temporally aligned with the heel strike and therefore does not constitute a reliable reference event.

Because heel-strike annotations and signal-derived peak positions do not coincide perfectly, a tolerance threshold was defined to account for minor temporal deviations. Based on typical step frequencies and the dataset’s sampling rate, a maximum difference of 123 milliseconds between the detected peak and the annotated ground-truth event was established as an acceptable margin of error.

#### Quality Management

To identify gait cycles unsuitable for inclusion in further analysis including the calculation of statistical features, a quality-assessment procedure was implemented. A mean gait cycle was computed for each recording, each participant, and each dataset, separately for each sensor location. In datasets where only a single recording was available per participant, the mean gait cycle at the recording and participant levels was identical. Cycles exceeding a predefined correlation threshold with their respective mean gait cycle were retained for further analysis, whereas those falling below the threshold were excluded. In this study, a correlation coefficient of 0.7 was selected as the cutoff threshold for inclusion.

#### Features

Ten statistical features that capture the regularity of the gait and its symmetry are calculated based on the gait cycles:

- mean value of dominant peaks (m_dp)
- standard deviation of dominant peaks (sd_dp)
- ratio of mean values of dominant and non-dominant peaks (r_m_dnp)
- ratio of standard deviations of dominant and non-dominant peaks (r_sd_dnp)
- mean standard deviation of dominant steps (m_sd_ds)
- ratio of mean standard deviations of dominant and non-dominant steps (r_m_sd_dns)
- standard deviation of standard deviations of dominant steps (sd_sd_ds)
- ratio of standard deviations of standard deviations of dominant and nondominant steps (r_sd_sd_dns)
- ratio of means of dominant and non-dominant step length (r_m_sl)
- ratio of standard deviations of dominant and non-dominant step length (r_sd_sl)

To obtain robust estimates of each feature for every recording, the largest possible number of subsets containing 10, 20, 30, 40, and 50 randomly selected gait cycles were generated, ensuring that each cycle was only used once. Feature values were first computed for each individual gait cycle and subsequently averaged within each subset (i.e., over 10, 20, 30, 40, or 50 cycles). The exemplary analyses reported in this work focus on features derived from subsets of 10 cycles as this subset size maximized the number of recordings eligible for inclusion.

In addition to these statistical features, the gait cycles uniformly resampled to 200 timepoints were incorporated as features for downstream analyses.

### Evaluation of the Harmonized Data

To assess the comparability of features across different recording conditions (here: dataset, i.e., sensor model and participants; body side; and sensor location) we conducted an exemplary set of pairwise comparisons (Table 2). Wherever feasible, all conditions except the one under investigation (highlighted in blue in Table 2) were held constant. This design allowed us to evaluate how features originating from similar contexts differ from each other, and how variations in dataset, body side, or sensor location influence both the extracted gait cycles and the derived statistical features.

**Table 2.**
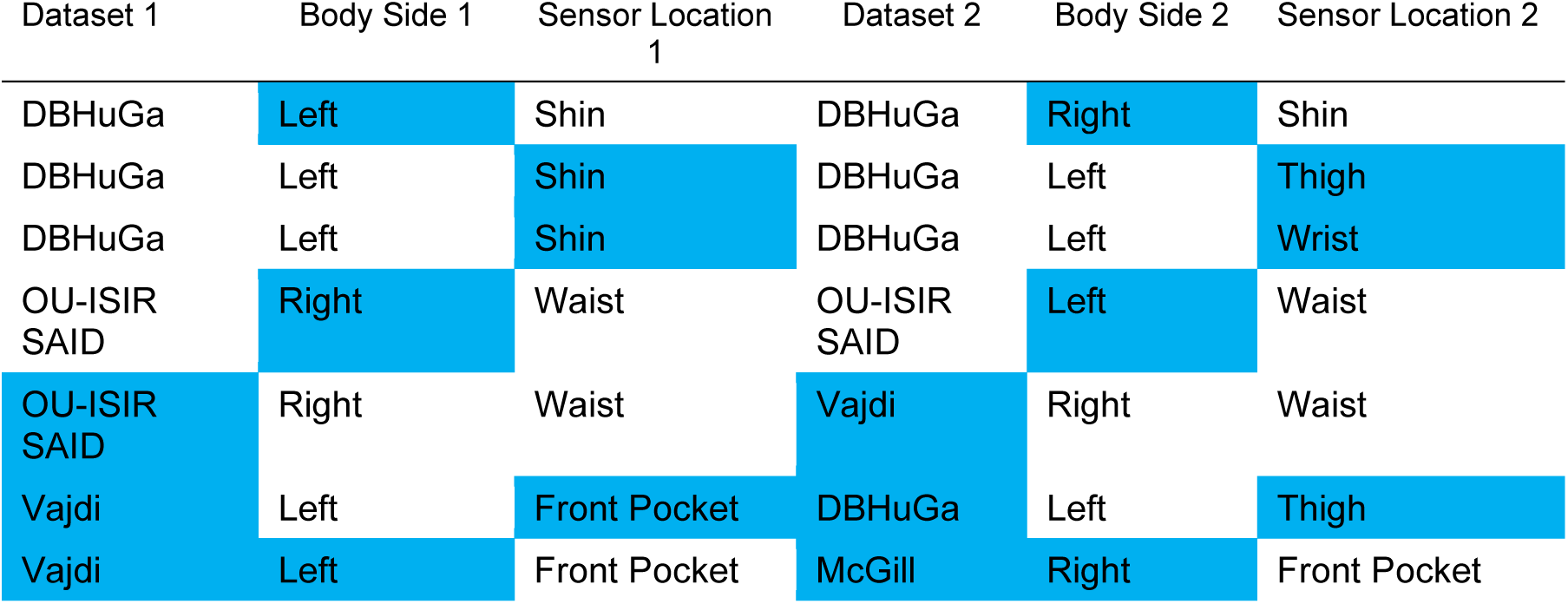
Dataset and sensor location pairs chosen for comparison. Overview of the chosen comparison for downstream analyses. Differences between the respectively selected pairs are marked blue.

When selecting sensor locations for comparison, the analyses were constrained by the number of usable features available for each location as well as the distribution of male and female participants across the relevant subsets of each dataset. These limitations excluded several comparisons that would have been of additional interest.

#### Statistical Comparisons

To obtain robust feature estimates for each sensor location and dataset, the individual values of the ten statistical features and the uniform gait cycles were averaged within each participant, body side, and sensor location. To assess differences in statistical features between pairs of conditions (Table 2), two-sided *t*-tests were conducted. Paired tests were applied when the compared conditions involved the same set of participants, unpaired tests otherwise. Because not all sensor locations yielded sufficient data to compute the full set of statistical features, some participants had to be excluded in the paired analyses to ensure that both samples comprised identical individuals.

It is important to note that statistical tests are intended solely as indicators of whether specific features may be unsuitable for harmonization; consequently, no correction for multiple comparisons was applied, instead, we report multiple α thresholds.

To evaluate the comparability of the uniform gait cycles, we computed Pearson correlation coefficients between all combinations of sensor locations and datasets. For each of those combinations we firstly calculate the average cycle per participant and then average those cycles for the further calculations. This analysis was performed separately for centered and non-centered sensor positions.

Furthermore, to characterize variability at the participant level, we conducted an analysis for each dataset–sensor-location combination. Specifically, we calculated the average correlation among the uniform gait cycles of different participants (across subject correlation) and compared these values with the correlations obtained among cycles belonging to the same participant (within subject correlation). This comparison provides insight into the extent to which gait-cycle representations are consistent within individuals relative to variability across individuals.

#### Classification of Gender

As a proof of concept for the data harmonization, we perform a gender-classification task, as gender information is available for most datasets included in the comparative analyses. Classification performance was evaluated for the same combinations of conditions used in the statistical comparisons (Table 2), enabling a direct assessment of how differences in dataset, sensor location, and body side affect predictions.

For the classification analyses, we used two machine learning algorithms: logistic regression as a linear classifier and random forests as a nonlinear classifier. The same two feature sets as in the statistical analysis were evaluated: (i) the ten statistical features and (ii) ten features derived from the uniform gait cycles (see below).

Six training–testing strategies were examined. These included training and testing on data from a single sensor; training and testing on combined data from both sensors, either preserving the dataset size of a single sensor or doubling it; and training on one sensor while testing on the other. For all six strategies, the amount of training data was matched to the size of the smaller dataset to ensure comparability across conditions. The data was sampled once in the beginning of the analysis, so all six strategies relied on the same participants.

To harmonize the dimensionality across feature sets, we used the first ten principal components derived from a principal component analysis (PCA) calculated on ten randomly chosen gait cycles per participant and condition.

Classification performance was evaluated using stratified cross-validation. To ensure statistical reliability, each fold contained a minimum of five samples, with the total number of folds capped at ten. For comparisons that yielded less than five folds we only report the balanced accuracy over all folds, for the remaining comparison we calculated mean and standard deviation across folds. Stratification was performed primarily with respect to gender, and in cases when data from two sources were combined also with respect to the originating dataset, to the extent permitted by the available sample distributions. To assess whether classification performance exceeded chance level, we conducted one-sided *t*-tests against chance level of 0.5. This test was skipped if the data was partitioned in less than five folds.

A conservative significance threshold of α = 0.01 was chosen to account for the multiple comparisons inherent in the evaluation across classifiers, feature sets, and training strategies. For completeness, results achieving only a threshold of α = 0.05 or even exceeding α = 0.001 are also indicated. In cases where the number of available folds was insufficient to support a *t*-test, we instead reported the weighted mean of the fold-wise accuracies as a descriptive summary measure.

To evaluate the influence of classification algorithm, feature set, and training strategy on model performance, we conducted mixed-effects analyses of variance (ANOVA) on the accuracy outcomes, with classification algorithm as within-subject factor and feature set as well as training-testing strategy as between-subject factors.

## Data Availability

No data was generated by this study. The following existing data sources were used: Human Gait Database The Human Gait Database (HuGaDB) is publicly available on Kaggle at https://www.kaggle.com/datasets/romanchereshnev/hugadb-human-gait-database. McGill University Gait Database The McGill University Gait Database (McGill) is publicly available at https://www.cs.mcgill.ca/∼jfrank8/data/gait-dataset.html. Movement Analysis in Real-world Environments using Accelerometers Gait Database The release agreement and terms of usage for the Movement Analysis in Real-world Environments using Accelerometers Gait Database (MAREA) are available at https://wiki.hh.se/caisr/index.php/Gait_database. Vajdi Gait Database The Vajdi Gait Database (Vajdi) is publicly available on Kaggle at https://www.kaggle.com/datasets/drdataboston/93-human-gait-database/data. OU-ISIR-Inertial Sensor Dataset The release agreement and download link for the OU-ISIR-Inertial Sensor Dataset (OU-ISIR-ISD) is available at http://www.am.sanken.osaka-u.ac.jp/BiometricDB/InertialGait.html. OU-ISIR-Similar Action Inertial Dataset The release agreement and download link for the OU-ISIR-Similar Action Sensor Dataset (OU-ISIR-SAID) is available at http://www.am.sanken.osaka-u.ac.jp/BiometricDB/SimilarActionsInertialDB.html. Database of Human Gait Performance The Database of Human Gait Performance (DBHuGa) dataset is publicly available at: https://doi.org/10.6084/m9.figshare.c.4892463. All technical artifacts including detailed technical documentation can be assessed via https://github.com/ReichenbachLabHHN/GaitDWH

http://www.am.sanken.osaka-u.ac.jp/BiometricDB/SimilarActionsInertialDB.html

http://www.am.sanken.osaka-u.ac.jp/BiometricDB/InertialGait.html

https://www.kaggle.com/datasets/drdataboston/93-human-gait-database/data

https://wiki.hh.se/caisr/index.php/Gait_database

https://www.cs.mcgill.ca/~jfrank8/data/gait-dataset.html

https://www.kaggle.com/datasets/romanchereshnev/hugadb-human-gait-database

## Acknowledgements

The striking image has been designed using resources from Flaticon.com.

## CReDiT statement (author contribution)

FC: Formal Analysis; Methodology; Software; Validation; Visualization; Writing – Original Draft Preparation; FS: Conceptualization; Data Curation; Formal Analysis; Methodology; Software; Validation; Writing – Original Draft Preparation; AR: Conceptualization; Formal Analysis; Project Administration; Supervision; Writing – Reviews & Editing.

## Data Availability

The datasets used in this study are publicly available. Detailed information on how to access the data can be found in the accompanying technical documentation (Supplementary Material S2).

